# Later-life Mortality and the Repeal of Federal Prohibition

**DOI:** 10.1101/2023.07.10.23292438

**Authors:** David S. Jacks, Krishna Pendakur, Hitoshi Shigeoka, Anthony Wray

**Affiliations:** National University of Singapore, CEPR, and NBER; Simon Fraser University; Simon Fraser University, University of Tokyo, IZA, and NBER; University of Southern Denmark

**Keywords:** Alcohol, federal prohibition, in utero exposure, later-life mortality

## Abstract

Despite a recent and dramatic re-evaluation of the health consequences of alcohol consumption, very little is known about the effects of in utero exposure to alcohol on long-run outcomes such as later-life mortality. Here, we investigate how state by year variation in alcohol control arising from the repeal of federal prohibition affects mortality for cohorts born in the 1930s. We find that individuals born in wet states experienced higher later-life mortality than individuals born in dry states, translating into a 3.3% increase in mortality rates between 1990 and 2004 for affected cohorts.

## 1. Introduction

While the harms of excessive alcohol consumption have long been understood (Rehm, 2011), only very recently has there been a dramatic re-evaluation of the health impacts of moderate levels of consumption. One of the starkest examples of this re-assessment comes from a Global Burden of Death, Injuries, and Risk Factors (GBD) study which concludes that the “level of [alcohol] consumption that minimizes health loss is zero” (GBD 2016 Alcohol Collaborators, 2018). Likewise, the personal and social harms of alcohol use have been ranked the highest among all drugs, including heroin and methamphetamine (Nutt, 2020). And this is not a story merely about the accumulation of academic and scientific evidence as consumers have responded in kind: worldwide, the share of active drinkers has been in decline since 2000 while per-capita use is declining in former strongholds of drinking activity and culture like the Anglosphere and Europe (World Health Organization, 2018).

The detrimental effects of in utero alcohol exposure on childhood development – first proposed by Lemoine *et al*. (1968) and definitively established by Clarren and Smith (1978) – has been amply demonstrated and is now widely appreciated. However, to our knowledge, there is no evidence on the later-life mortality effects of in utero exposure to alcohol. And while there is a well-established literature in economics considering quasi-experiments that substantially eased or restricted access to alcohol, these policy changes are oftentimes short-lived, limited in their geographic scope, or too recent in the past to speak to the effects of alcohol exposure on later-life mortality (Aizer and Currie, 2014; Almond, Currie, and Duque, 2018; Carpenter and Dobkin, 2009, 2011; Kueng and Yakovlev, 2021; Nilsson, 2017).

This paper addresses these issues head on and fills the gap in our understanding by assessing the long-run effects of federal prohibition’s repeal in the 1930s on later-life mortality and offering up potential physiological mechanisms for the same. In so doing, we confront a common misunderstanding about the nature of federal prohibition: there was no uniform policy change with restrictions on alcohol “turning off” precisely in December 1933 when federal prohibition was repealed. Indeed, the decentralized nature of American government and the political concessions necessary to bring about repeal worked to ensure that there was ample geographic and temporal heterogeneity in restrictions on alcohol well after federal prohibition ended.

Our goal is to identify the causal effects of in utero exposure to alcohol on later-life mortality. To do so, we use data on state by year variation in alcohol prohibition coupled with annual death rates from 1990 to 2004 for cohorts born in the 1930s. We conduct event-study analysis in the context of a difference-in-differences research design that exploits the staggered timing of federal prohibition’s repeal. We find evidence that allowing for legal alcohol sales (that is, transitioning from “dry” to “wet” status) at the state level is associated with a 3.3% increase in mortality rates between 1990 and 2004. Critically, we show that these effects are localized to the in utero period as cohorts which were already in early childhood when states became wet do not see equivalent increases in later-life mortality.

We also speak to the potential physiological mechanisms underlying this result by examining repeal’s effect on cause-specific mortality rates. We find that in utero exposure to alcohol availability is associated with increases in mortality rates for heart disease and stroke in later life. We also have good reasons to believe that these results are not spurious: we find no corresponding increase in later-life mortality arising from motor vehicle accidents, another leading cause of death which is plausibly unrelated to in utero alcohol exposure. We furthermore demonstrate that our results are unaffected when we control for exposure to both the Great Depression and New Deal spending. This suggests that our results are not driven by other confounding events which may be correlated with the timing of prohibition’s repeal across states and years. Finally, we examine potential heterogeneity along the lines of sex and race, finding that females and males as well as non-white and white people are similarly affected, suggesting our baseline results are not driven by other mechanisms such as differential healthcare access.

This paper contributes to the literature in two ways. First, we present the first causal evidence that the well-known, determinantal effects of in utero exposure to alcohol also extend to mortality in later life. Prior work has documented in utero effects of potential alcohol exposure on contemporaneous infant mortality (Jacks, Pendakur, and Shigeoka, 2021) and subsequent labor market outcomes (Nilsson, 2017), but not on outcomes observed in later life. Second, we add to a burgeoning literature on the consequences for later-life mortality of economic shocks and policy interventions occurring in the United States during the 1930s. Aizer *et al*. (2016) find that the Mothers’ Pension program increased longevity for male children of recipients. Duque and Schmitz (2023) find that children most exposed to the deprivations of the Great Depression experienced higher mortality in later life, a result which is likely driven by the Great Depression’s effect on accelerated epigenetic aging (Schmitz and Duque, 2022). At the same time, Jou and Morgan (2023) find that after accounting for the endogeneity of New Deal spending, the latter moderated – and in some instances fully reversed – the negative effects of the Great Depression.

Understanding the effects of repeal might also be important with respect to contemporary policy issues related to alcohol. The US Surgeon General’s initial warning about the risks associated with alcohol consumption during pregnancy was issued in 1981. But in the 1930s, the general public had little definitive knowledge of the potential negative effects of alcohol consumption during pregnancy on child development (Warner and Rosett, 1975). Thus, our estimates are potentially not driven by differences in avoidance behaviours by mothers of different socioeconomic status. Finally, we note that the scope for policy interventions is still large. Although information about the risks associated with alcohol consumption during pregnancy is now widely understood, in the United States, over 50% of women of childbearing age drink (Tan *et al*., 2015) while an estimated 15% of women continue to drink during pregnancy (Popova *et al*., 2015) and up to 9% of all children suffer from some form of fetal alcohol spectrum disorders (May *et al*., 2018).

## 2. Historical background

A long-standing temperance movement in the United States — and indeed globally (Schrad, 2021) — quickly culminated with the federal prohibition of alcohol in 1920. The US Senate first proposed a constitutional amendment in December 1917, and by January 1919, the 18^th^ Amendment was ratified with the country becoming dry on January 17, 1920. Here, we briefly discuss the features of federal prohibition and its repeal that are important for our identifying variation.

First, federal prohibition appealed to a very wide range of the public and was surprisingly effective. Agitation for federal prohibition was supported by a remarkably wide range of interests — patriotism, progressivism, religion, and women’s rights among others (Rorabaugh, 2018). Widespread support for prohibition also translated into a decline in consumption. As can be seen in Panel A of Figure 1, in the first year of repeal (1934), apparent per capita alcohol consumption was 63% lower than its pre-prohibition peak in 1910.^1^ What is more, the shock of prohibition apparently lingered in the consumption habits of Americans as it took at least until the 1970s for per-capita alcohol consumption to surpass its previous heights. While the passage of the 18^th^ Amendment entailed a near-complete prohibition on the production, sale, and transportation of alcohol, it did not ban individual consumption and possession of alcohol. These were rather subject to varying degrees of restriction at the city, county, and state level. Instead, prohibition is best thought of as a substantial tax on alcohol which served to reduce consumption (Asbury, 1950; Cook, 2007).

**Figure 1:**
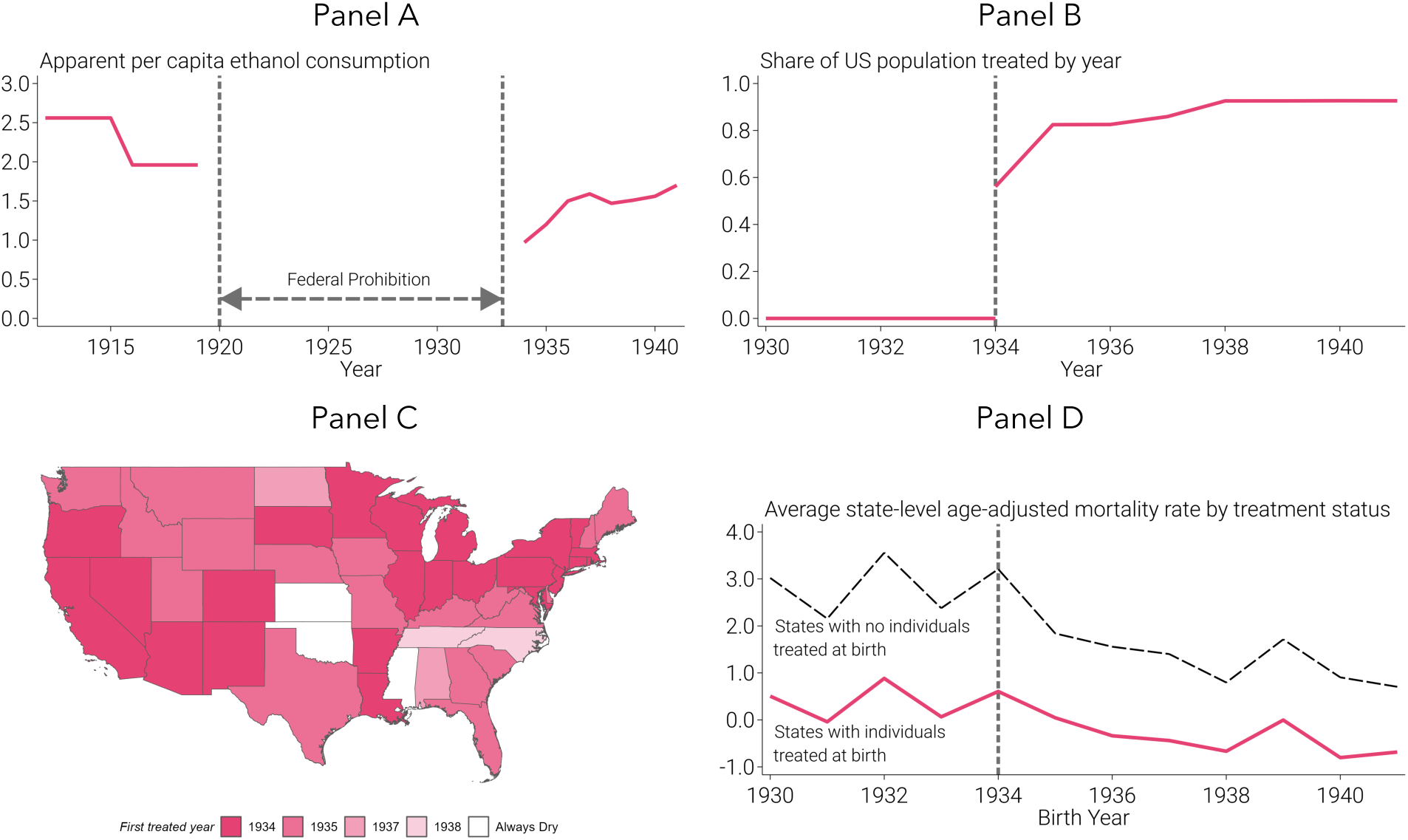
Alcohol Consumption, Prohibition Status, and Age-adjusted Mortality. Panel A depicts apparent alcohol consumption on a per capita basis which is derived from alcoholic beverage sales data and measured in gallons of pure ethanol. Source: LaVallee and Yi (2011). Panel B uses the prohibition status of all US counties (n = 3,111) using population weights derived from the US Census. In our baseline specification, wet states are those which allow for (at least some) alcohol sales anywhere within their borders. Source: Jacks, Pendakur, and Shigeoka (2021). Panel C displays the initial year when a state turned wet. The only states that remained “always dry” from 1930 to 1941 form the control group (i.e., Kansas, Mississippi, and Oklahoma). Map source: Manson *et al*. (2022). Panel D plots averages of residuals from a regression of later-life mortality rates per 1,000 population on fixed effects for age at death, separately for states that ever turn wet between 1930 and 1941 (solid red line) and the three states in the control group that remained “always dry” (dashed black line).

Second, the process of federal repeal was also remarkably quick, implying the absence of anticipation. Concerns over the new reach of the federal government and perceptions of rising criminal activity rose throughout the 1920s (Asbury, 1950; Garcia-Jimeno, 2016; Okrent, 2010) which were accentuated by the onset of the Great Depression. Given the dire fiscal straits of the early 1930s (Blocker, 2006; Rorabaugh, 2018), various levels of government increasingly eyed the return of alcohol sales as a potential source of revenue. On March 22, 1933, Roosevelt started this process by amending the National Prohibition Act, allowing for the production and sale of low-alcohol beer. Within the year, the 21^st^ Amendment was ratified by special conventions in 38 states, thereby repealing the 18^th^ Amendment and ending federal prohibition on December 5, 1933.

Finally, the chief compromise for achieving ratification of the 21^st^ Amendment came in allowing for local option elections to determine liquor laws deemed appropriate for local conditions (Kyvig, 2000).^2^ This compromise ensured that the process of repeal was not uniform across states, affording us an important source of variation in prohibition status which we exploit below. As states quickly and sometimes unexpectedly opted for repeal of federal prohibition, they reverted to the *status quo ante* established by any legislation related to alcohol control that pre-dated federal prohibition. Many jurisdictions that wanted changes in their respective prohibition status, thus, had to wait for the arrival and passage of enabling legislation. The patchwork regulatory regime that emerged immediately after repeal worked to ensure that the timing of such transitions was a function of idiosyncratic local factors (Childs, 1947; Clark, 1965; Fosdick and Scott, 1933; Harrison and Laine, 1936). That is, they were likely uncorrelated with other potential policy changes (Jacks, Pendakur, and Shigeoka, 2023).^3^

## 3. Data

Our data are drawn from two main sources: annual counts of death by state of birth and year of birth have been extracted from the Multiple Cause-of-Death Mortality database and the US Census is used to calculate the size of surviving cohorts in 1990 while annual, indicators of state-level prohibition status have been constructed from contemporary sources. These are discussed along with details of data construction in sections 3.1 and 3.2, respectively.

### 3.1 Dependent variable: mortality and surviving cohort size

We consider the universe of deaths in the US from 1990 to 2004 taken from the Multiple Cause-of-Death Mortality database (National Center for Health Statistics, 1990— 2004).^4^ The underlying data are based on death certificates for all US residents. These certificates contain a primary cause of death and up to twenty additional multiple causes along with some limited demographic information (principally, the perceived race and sex of the deceased). Critically for our purposes, they also contain information on not only individuals’ death date and place of death but also their birth date and place of birth. Given the timing of events related to federal prohibition and its repeal, we focus on those cohorts born from 1930 to 1941. We collapse these data to obtain the number of deaths (*D_bst_*) in year (*t*) for a given state of birth (*s*) by year of birth (*b*) cohort.

We also estimate the surviving cohort size in 1990 (*A_bs1990_*) from the 5% sample of the 1990 Census from IPUMS (Ruggles *et al*., 2023). We multiply observations in the 1990 Census by the corresponding sample weights and collapse the data for state of birth by year of birth cohorts. We then iteratively compute *A_bst_* for *t* = 1991, …, 2004 by subtracting annual deaths, *D_bst_*. We combine the counts of deaths (*D_bst_*) and the surviving cohort size (*A_bst_*) to construct annual mortality rates, *d_bst_* = 1000 * *D_bst_* / *A_bst,_* per 1,000 individuals. Thus, our variable of interest is the mortality rate in year (*t*) for a given state of birth (*s*) by year of birth (*b*) cohort.

### 3.2 Treatment variable: prohibition status

Here, we build on previous data collection efforts. Jacks, Pendakur, and Shigeoka (2021) reconstructs the prohibition status of all US counties for the key post-repeal period from 1934 to 1941 using an array of sources (Culver and Thomas, 1940; Distilled Spirits Institute, 1935, 1941; Harrison, 1938; Thomas and Culver, 1940). The underlying dataset relies on the sharpest distinction in prohibition status available at the county level: dry versus wet. That is, it allows for comparisons across jurisdictions for which no alcohol sales are permitted (dry) to those for which at least some alcohol sales are permitted (wet). Panel B of Figure 1 demonstrates that the transition away from federal prohibition was swift as 80% of the US population found itself living in newly wet jurisdictions by 1935. This process continued — albeit more slowly — in the remainder of the 1930s with this share reaching nearly 85% in 1938. In contrast, there was virtually no change in this share from that point forward.

In order to match annual all-cause mortality rates by state of birth (*s*) discussed in section 3.1, we convert this binary measure of county-level wet status into a state-level indicator for whether *any* county in a state goes wet.^5^ We do so for two reasons: (1) this approach leans towards producing more conservative estimates of repeal’s effect on later-life mortality; (2) this approach partially controls for cross-county (albeit intra-state) externalities arising from uncoordinated changes in prohibition status as in Jacks, Pendakur, and Shigeoka (2021). In robustness exercises below, we also use other indicators capturing whether an entire state turns wet via statewide legislation or whether at least 50% of a state’s population resided in counties that allowed alcohol sales with qualitatively the same results.

We can also consider the spatial distribution of dry and wet states by year through 1931 as in Panel C of Figure 1. By 1936, the remaining hold-out states for prohibition were along the central axis of the US (Kansas, North Dakota, and Oklahoma) along with large parts of the Southeast (Alabama, Mississippi, North Carolina, and Tennessee). However, this core of dry states was whittled away through time: Alabama and North Dakota jettisoned statewide prohibitions in 1937 as did North Carolina and Tennessee in 1938, leaving Kansas, Mississippi, and Oklahoma as the only “always dry” states and, thus, forming our primary control group.

## 4. Empirical framework

For our baseline results, we first estimate a set of event studies that exploits variation in the exposure to wet status coming from two sources: the timing of birth for individuals and the timing of repeal across states. In particular, we estimate the following:

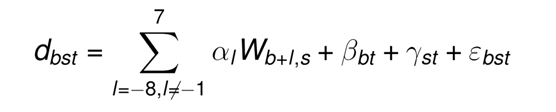

where *b* indexes year of birth (12 years from 1930 to 1941), *s* indexes state of birth (48 states), and *t* indexes observed years of deaths (15 years from 1990 to 2004). Implicitly, the age range of mortality we examine is from 49 to 74 years as we observe those that were born in the period from 1930 to 1941 in a window of mortality from 1990 to 2004. Due to the restricted window of mortality we have at our disposal, the age ranges at which we observe *d_bst_* are not the same across cohorts born in different years.

We define wet status *W_b+l,s_* as a binary treatment where *W_b+l,s_* = 1 indicates that a state was at least partially wet in the *l*’th lead or lag of the year of birth (b). That is, *l* = 0 represents a cohort born in the same year as when their state of birth transitioned to wet status. Values of *l* less than 0 indicate that a cohort was born before their state of birth became wet: *l* = -2, for example, indicates that a cohort is two years of age when their state of birth’s transition to wet status occurred. Similarly, values of *l* greater than 0 indicate that they were born after their state of birth became wet: *l* = 2, for example, indicates that a cohort was born two years after their state of birth’s transition to wet status occurred. We use the normalization that *α _-1_* = 0, meaning that all reported treatment effects are relative to the year before birth.^6^

Our regression model also includes two sets of interacted fixed effects as the range of ages at which we observe deaths varies for each birth cohort. First, we add year of birth by observed year of death fixed effects (*ý _bt_*) for the 12 years of birth interacted with the 15 observed years of death to flexibly control for differential trends in life expectancy across cohorts, e.g., a fixed effect for deaths in 1990 of those born in 1930. Second, we add state of birth by observed year of death fixed effects (*ψ _st_*) for the 48 states interacted with the 15 observed years of death to flexibly control for differential trends in life expectancy across states, e.g., a fixed effect for deaths in 1990 of those born in Texas. We note that this specification is more flexible than simply including separate fixed effects for year of birth (*b*), state of birth (*s*), and observed year of death (*t*) as is seen in some of the related literature.

A recent literature argues that the standard OLS two-way fixed effects (TWFE) estimator can be biased in panel-data settings with staggered adoption like ours if there is heterogeneity in the treatment effect across cohorts and/or over time. Here, we use the estimator proposed by Sun and Abraham (2021) that only uses never-treated units as controls.^7^For the sake of comparison, we also report OLS estimates of the dynamic TWFE model in our event studies.

All regressions are weighted by the surviving cohort size for each state of birth by year of birth by observed year of death cell, e.g., those who were born in Texas in 1930 and die in 1990. Likewise, all standard errors are clustered at the state of birth by year of birth level to account for potential within state of birth by year of birth serial correlation of arbitrary form.

## 5. Main results and robustness

We begin with a presentation of underlying trends in later-life mortality rates for treated versus untreated states. We start by taking the residuals from an OLS regression of annual all-cause mortality rates solely on fixed effects for age at death. We partial out the age-at-death fixed effects rather than plot the raw data as the range of ages at which we observe deaths for each birth cohort varies. Panel D of Figure 1 plots the average value of these residuals separately for states that ever turn wet between 1930 and 1941 (depicted by the solid red line) and the three states in the control group that remained “always dry” (depicted by the dashed black line). Prior to repeal in late 1933, states with no (future) individuals treated at birth register higher age-adjusted mortality rates, but importantly the two series move in parallel, suggesting that the common trends assumption is satisfied. From 1933, both series begin to decline, but the gap narrows. This suggests that while later-life mortality rates declined for all cohorts born after 1933, they apparently did more so for those born in “always dry” states. The empirical framework outlined in section 4 allows us to more rigorously assess this possibility with the inclusion of a large battery of fixed effects.

Our results are presented in three parts: first, we consider our main event studies for later-life all-cause mortality rates and then later-life cause-specific mortality rates to reveal potential physiological mechanisms; second, we supplement these event studies with aggregated coefficient estimates to aid the interpretation of magnitudes as well as demonstrate that our results are robust to the inclusion of control variables and alternative specifications; and finally, we explore potential heterogeneity in our main results along the lines of sex and race.

### 5.1 Event studies for annual mortality rates

Here, we report the results of various event study analyses as outlined in section 4. We start with event studies that exploit the variation in the exposure to wet status coming from the timing of birth for individuals and the timing of repeal across states. Figure 2 presents OLS estimates of the event-study and the Sun and Abraham (2021) bias-corrected estimates of the same model using the three never-treated states of Kansas, Mississippi, and Oklahoma as the control group. We note that these estimates are very similar to each other, suggesting that the bias in the OLS estimates is small. In light of this, we exclusively refer to the Sun and Abraham bias-corrected estimates in the following discussion.

**Figure 2:**
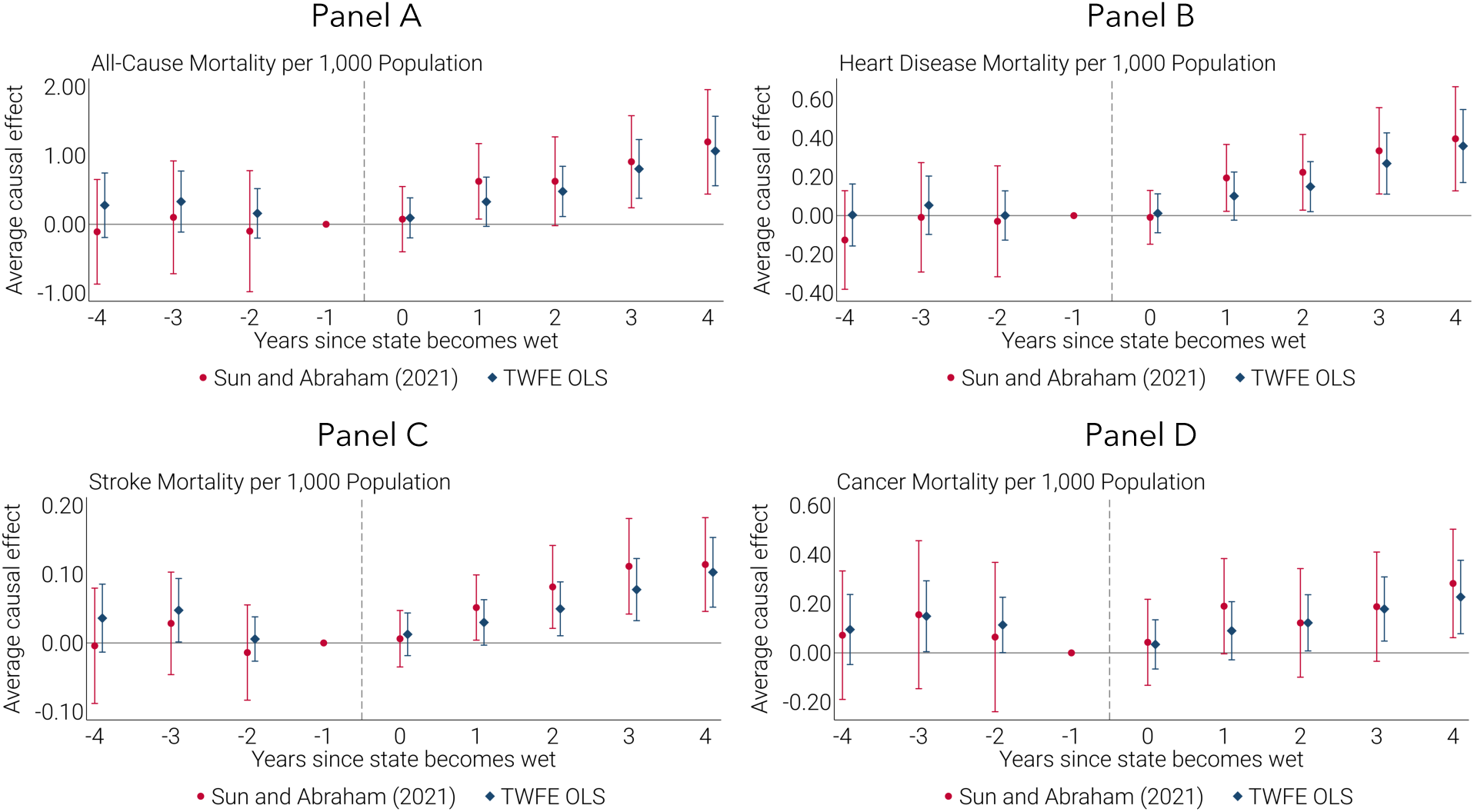
Event Studies on All-Cause and Cause-Specific Mortality. Each panel displays event studies with all-cause mortality (panel A) or cause-specific mortality per 1,000 population as the dependent variable (heart disease in panel B, stroke in panel C, and cancer in panel D). The unit of observation is the annual mortality rate for a given state of birth by year of birth cohort. The sample includes the 1930 to 1941 birth cohorts. Mortality is observed between 1990 and 2004. Each panel shows event studies based on two estimation methods: Sun and Abraham (2021) and TWFE OLS. The coefficients for event-time periods *l* = −8 to *l* = −5 and *l* = 5 to *l* = 7 are omitted for clarity and due to compositional changes in the set of treated states. All specifications include the year of birth by death year and state of birth by death year fixed effects. Observations are weighted by the surviving cohort size for each state of birth by year of birth by death year cell. Standard errors are clustered at the state of birth by year of birth level.

The results of this exercise for all-cause mortality are depicted in Panel A of Figure 2. Naturally, in the case of potential in utero effects, we expect to see non-zero effects emerge after treatment but not before, provided that the treatment is not confounded with existing pre-trends. The point estimates for *l* > 0 are all positive and statistically significant. These results suggest that the potential availability of alcohol for parents before birth drives differential later-life mortality across dry and wet states. They also suggest that we should consider physiological mechanisms potentially related to in utero maternal alcohol consumption.

With respect to the fairly consistent null results attached to *l* = 0 in Figure 2, we note that we only observe legal prohibition status at an annual frequency and, thus, there is uncertainty about when in a particular year the change in status occurred. Moreover, even in the case where a precise date of status change is known, there are likely to be lagged effects due to discrepancies in the timing between when changes in legislation occurred and when they became effective (which were often significant) and between when changes became effective and when retail outlets for legal alcohol were established.

In contrast, the point estimates for *l* < -1 are nearly zero and statistically insignificant. Therefore, these results do not suggest that there is a discernible pre-trend or that the potential availability of alcohol for parents after birth (ages one and on) drives differential later-life mortality across “always dry” and wet states. We can then exclude potential mechanisms related to post-natal parental investment or domestic violence as these would suggest an effect for birth cohorts born prior to a state going wet (e.g., *l* = -2) as well.

We next speak to the potential physiological mechanisms underlying this result by examining cause-specific mortality. In particular, we examine, in turn, later-life mortality rates coming from heart disease, stroke, and cancer, which were the three leading causes of death in 2004 (Centers for Disease Control and Prevention, 2007).^8^ Panels B, C, and D depict the respective event studies for these causes of death. For heart disease and stroke, there is a clear resemblance between their profile and that for all-cause mortality: the estimates suggest that there are no discernible pre-trends, there are null results in the year after states transition to wet status, and there are significant increases in later-life mortality rates from that point forward. The evidence for mortality from cancer in later-life is more muted as the point estimates are less precise, both before and after states transition to wet status.

We also have good reasons to believe that these results are not spurious: a fully equivalent event study for motor vehicle accidents (the fifth leading cause of death in 2004) yields point estimates with no discernible trend before or after repeal and which are grossly insignificant (see Figure A2 in Appendix A). This placebo test suggests that our results on later-life mortality are not driven by other confounding events which are correlated with the timing of prohibition’s repeal across states and years.

In the next section, we aggregate the event study estimates. First, we do so to provide a single point estimate which summarizes our event studies and illustrates the magnitude of our results. Second, we also demonstrate the robustness of our main results to the inclusion of various controls and the use of various specifications.

### 5.2 Aggregate event study estimates

Having shown that there were no discernible pre-trends in the all-cause mortality rate, we turn to a consideration of the aggregation of the event study coefficients from Figure 2. In Table 1, we report the average effect over the first five years of repeal (from *l* = 0 to *l* = 4) which matches the set of post-repeal coefficients reported in Figure 2. Thus, it reports a summary measure of repeal’s effect on later-life mortality. The specification for Column 1 corresponds to the event study in Panel A of Figure 2, where all-cause mortality rates are a function of the state of birth by death year and year of birth by death year fixed effects. We find a statistically significant (at the 1% level) increase in later-life all-cause mortality rates by 0.69 deaths per 1,000 population for those cohorts who were born in a wet state within five years of repeal. One way of putting the magnitude of this result into context is by comparing it to the underlying all-cause mortality rate for not-yet-treated cohorts which was 18.85 deaths per 1,000. Thus, repeal was associated with a 3.6% (=0.69/18.85) increase in later-life mortality rates between 1990 and 2004.

**Table 1:** Aggregate Event Study Estimates for All-Cause Mortality.

|  | (1) | (2) | (3) | (4) | (5) | (6) |
| --- | --- | --- | --- | --- | --- | --- |
|  | Baseline<br>without<br>controls | Baseline<br>with<br>controls | Last<br>treated | State level<br>transitions | Majority<br>transitions | State level<br>clustering |
| Wet status (=1) | 0.69<br>(0.25)<br>[2.73] | 0.63<br>(0.29)<br>[2.18] | 0.62<br>(0.24)<br>[2.53] | 0.97<br>(0.30)<br>[3.27] | 0.66<br>(0.18)<br>[3.64] | 0.63<br>(0.26)<br>[2.45] |
| Mean of Y | 18.85 | 18.85 | 18.85 | 17.57 | 18.56 | 18.85 |
| Percent effect relative to mean | 3.6 | 3.3 | 3.3 | 5.5 | 3.6 | 3.3 |
| Observations | 8,640 | 8,640 | 5,760 | 8,640 | 8,640 | 8,640 |
| Year of birth by death year FEs | X | X | X | X | X | X |
| State of birth by death year FEs | X | X | X | X | X | X |
| State by year of birth controls |  | X | X | X | X | X |

The remainder of Table 1 establishes the robustness of our results to various other specifications.^9^ Column 2 replicates the main specification from Figure 2 but includes two critical control variables: one for the log of real New Deal spending per capita and another for the log of real disposable personal income per capita, both measured at the state level and drawn from Fishback (2015) and the Bureau of Economic Analysis (2023), respectively. Together, these variables are intended to control for the combined effects of the Great Depression and the related policy response coming from the New Deal which have been shown to have influenced later-life mortality in the work of Duque and Schmitz (2023) as well as Jou and Morgan (2022). The concern here is that a state’s prohibition status may have been related to both the severity of the economic downturn as well as the extent of government relief. In any case, we are reassured that our estimate of repeal’s effect remains virtually unchanged with the inclusion of these controls as the associated estimated coefficient and percent effect now register at 0.63 deaths per 1,000 and 3.3%, respectively.

Columns 3 through 6 also retain these controls and instead consider different definitions of the treatment as well as a different level of clustering. First, we address concerns that the never-treated states followed different trends and, thus, are not an appropriate control group. Column 3 uses the states in the last treated group as the control. Since these states (North Carolina and Tennessee) transitioned to wet status in 1938, this specification necessarily drops observations for 1938 to 1941. In any case, the estimate is hardly changed.

Next, we contend with the fact that, in some states, only a portion of the population was exposed to the repeal of prohibition which would tend to bias our estimates downward. In Column 4, a state is considered to be treated only if the entire state turns wet via statewide legislation while in Column 5 a state is considered to be treated if at least 50% of its population resided in a county that allowed alcohol sales. As might be expected, all these specifications yield larger (and statistically significant) coefficients, ranging from 0.66 to 0.97 per 1,000 individuals. Consequently, the estimated percent effects ranges from 3.6% to 5.5%. Lastly, Column 6 reverts to the original definition of the treatment variable and specification as in Column 2 but allows for standard errors to be clustered more conservatively at the state of birth level. In this instance, the results remain statistically significant at the 5% level.

Cause-specific mortality results are reported in Table 2 and follow our baseline specification inclusive of all controls and fixed effects as in Column 2 of Table 1. Column 1 reproduces the baseline estimate for all-cause mortality. Columns 2 through 4 consider the same specification and report the equivalent results for cause-specific mortality rates from heart disease, stroke, and cancer, respectively. Given our previous event studies, this exercise unsurprisingly yields uniformly positive and significant coefficients for repeal’s effect on heart disease and stroke. The estimated percent effects are sizeable, ranging from 4.3% for heart disease and 7.2% for stroke. In contrast, the estimate for cancer is much less precisely estimated and not statistically significant at conventional levels.

**Table 2:** Aggregate Event Study Estimates for Cause-Specific Mortality.

|  | (1) | (2) | (3) | (4) |
| --- | --- | --- | --- | --- |
|  | All-cause<br>mortality | Heart<br>disease | Stroke | Cancer |
| Wet status (=1) | 0.63<br>(0.29)<br>[2.18] | 0.22<br>(0.08)<br>[2.92] | 0.06<br>(0.03)<br>[2.41] | 0.16<br>(0.10)<br>[1.63] |
| Mean of Y | 18.85 | 5.12 | 0.88 | 6.72 |
| Percent effect relative to mean | 3.3 | 4.3 | 7.2 | 2.3 |
| Observations | 8,640 | 8,640 | 8,640 | 8,640 |
| Year of birth by death year FEs | X | X | X | X |
| State of birth by death year FEs | X | X | X | X |
| State by year of birth controls | X | X | X | X |

To round out our consideration of robustness, Appendix B reports results from when we extend our measure of all-cause mortality rates back to 1979. Appendix C reports estimates from using a continuous treatment, i.e., exploiting the share of a state’s population living in wet counties. Appendix D reports the results when we use cumulative all-cause mortality rates over all observed years of death from 1990 to 2004, rather than annual all-cause mortality rates as in our baseline specification. We are reassured that we find materially the same results throughout these exercises.

### 5.3 Heterogeneity analysis

Finally, we can also consider the possibility that repeal differentially affected all-cause mortality rates on the basis of race and sex. Starting with heterogeneity on the basis of sex, Columns 1 and 2 of Table 3 report the aggregate event study estimates for females and males, respectively. The point estimate for females is 0.45 and statistically significant while that for males is 0.91 but not statistically significant. Consequently, the two point estimates are not statistically distinguishable from one another.

**Table 3:** Aggregate Event Study Estimates for Heterogeneity by Sex and Race.

|  | (1) | (2) | (3) | (4) |
| --- | --- | --- | --- | --- |
|  | Females | Males | Non-white | White |
| Wet status (=1) | 0.45<br>(0.19)<br>[2.37] | 0.91<br>(0.55)<br>[1.65] | 0.91<br>(0.88)<br>[1.04] | 0.44<br>(0.29)<br>[1.53] |
| Mean of Y | 14.74 | 23.78 | 25.20 | 18.45 |
| Percent effect relative to mean | 3.0 | 3.8 | 3.6 | 2.4 |
| Observations | 8,640 | 8,640 | 8,560 | 8,640 |
| Year of birth by death year FEs | X | X | X | X |
| State of birth by death year FEs | X | X | X | X |
| State by year of birth controls | X | X | X | X |

Columns 3 and 4 report the equivalent results for all-cause mortality rates on the basis of race. The point estimate for non-white people is 0.91 while that for white people is 0.44. However, neither are statistically significant. We suspect that part of the greater imprecision for non-white people is driven by noise in the count of non-white people in the 1990 Census for certain states and years of birth and/or the assignment of race upon death (see Appendix E for the corresponding event studies). In any case, these results suggest that our baseline results are not driven by other mechanisms such as differential healthcare access along the lines of sex or race.

## 6. Conclusion

We find evidence that allowing for legal alcohol sales at the state level is associated with a 3.3% increase in mortality rates between 1990 and 2004 for cohorts born in the 1930s when federal prohibition was repealed. To our knowledge, this is the first evidence on the effects of in utero exposure to alcohol on later-life mortality. To the extent that the weakest of embryos are eliminated via a culling effect (Jacks, Pendakur, and Shigeoka, 2021), the scarring effect of in utero alcohol exposure to alcohol on later-life mortality estimated in this paper may actually prove to be a lower bound.

And while we do not observe actual consumption of alcohol during pregnancy by mothers (that is, the critical “first stage”), our results collectively point toward maternal alcohol consumption as the underlying mechanism: we show that these effects are localized to the in utero period as cohorts which were already in early childhood when states became wet do not see equivalent increases in later-life mortality; we do not find effects for sources of later-life mortality such as motor vehicle accidents which are plausibly unrelated to in utero exposure; and we find relatively uniform effects across sex and race.

## Data Availability

The mortality records used here are publically available.
The Prohibition status data is manually collected and thus available upon request.

## Footnotes

* We appreciate feedback from Simon Woodcock as well as seminar and conference participants at the National University of Singapore, the 2021 Asian and Australasian Society of Labour Economics Conference, the 2022 Canadian Economics Association, and 2022 CEPR Economic History Symposium meetings. We also gratefully acknowledge research support from the National University of Singapore and the Social Sciences and Humanities Research Council of Canada. Jacks: National University of Singapore, CEPR, and NBER; Pendakur: Simon Fraser University; Shigeoka: Simon Fraser University, University of Tokyo, IZA, and NBER; Wray: University of Southern Denmark.

1 Of course, the first year of repeal corresponds with the first full year of recovery from the Great Depression, so some of this 63% decline might also be attributable to economic conditions and not federal prohibition per se. Cigarettes were another stimulant on which discretionary income could be spent, but for which there was no prohibition: from 1929 to 1934, per-capita cigarette consumption fell by roughly 10% (Warner, 1985). This suggests that federal prohibition’s likely effect on individual’s alcohol consumption was indeed quite large.

2 These elections have a long standing in American history and give the electorate the right to vote on liquor control by referendum. That is, local majority preferences determine whether a jurisdiction allows for or prohibits the sale of alcohol. Many states opted out from local option elections entirely while others allowed for referenda to be periodically held.

3 Furthermore, Appendix D of Jacks, Pendakur, and Shigeoka (2021) considers the possibility that individuals may have migrated to counties in response to their respective maintenance or repeal of prohibition at the local level. Analysis of county-level measures of net migration in 1940 finds no relationship between changes in prohibition status and county-level changes in population. Thus, there is little evidence to the effect that changes in prohibition status drove intercounty – and presumably, interstate – migration patterns in this period.

4 We use deaths from 1990 in our main specifications instead of 1979, the first year for which state of birth is available in the MCOD database. We do so in order to minimize any measurement error stemming from having to construct the size of surviving cohorts (*Abst* below) from the Census. 1990 as a starting point also seems apt for the fact that we are interested in later-life mortality here. In any case, our results are robust to using either 1979 or 1990 as the earliest sample cohort, as shown in Appendix B. Finally, we are limited to using deaths until 2004 as there is no information on individuals’ state of birth after that year in the unrestricted version of the MCOD database.

5 Recently, estimators have been proposed for settings like these with staggered treatment timing, but they do so in the context of a binary treatment that stays on after adoption (Callaway and Sant’Anna, 2021; de Chaisemartin and Haultfoeuille, 2020; Sun and Abraham, 2021). While these estimators are robust to treatment effect heterogeneity, the latter can make it difficult to interpret treatment parameters across different values of the treatment in the continuous treatment setting (Callaway, Goodman-Bacon, and Sant’Anna, 2021). Nonetheless, Appendix C reports OLS estimates using a treatment continuously defined on the share of a state’s population that resides in a wet county, yielding qualitatively similar results.

6 See Figure A1 in Appendix A for the distribution of treated states by event time period.

7 Among recently proposed estimators in the context of staggered treatments, to our knowledge, only that of Sun and Abraham (2021) allows us to flexibly control for two sets of interacted fixed effects. All other estimators would only allow us to include state of birth and year of birth fixed effects. In later robustness exercises, we use cumulative all-cause mortality rates over all observed years of death from 1990 to 2004, rather than annual all-cause mortality rates as in our baseline specification. Since we only include state of birth and year of birth fixed effects in this specification, we also report the results coming from the estimators of Callaway and Sant’Anna (2021) and de Chaisemartin and D’Haultfoeuille (2020) with materially the same results.

8 See Table A1 in Appendix A for the crosswalk of ICD9 and ICD10 codes used to construct cause-specific mortality rates

9 Appendix Figure A3 presents the event studies that underlie the aggregated coefficient estimates.

## Online Appendix for “Later-life Mortality and the Repeal of Federal Prohibition”

### Appendix A: Additional Figures and Tables

**Figure A1:**
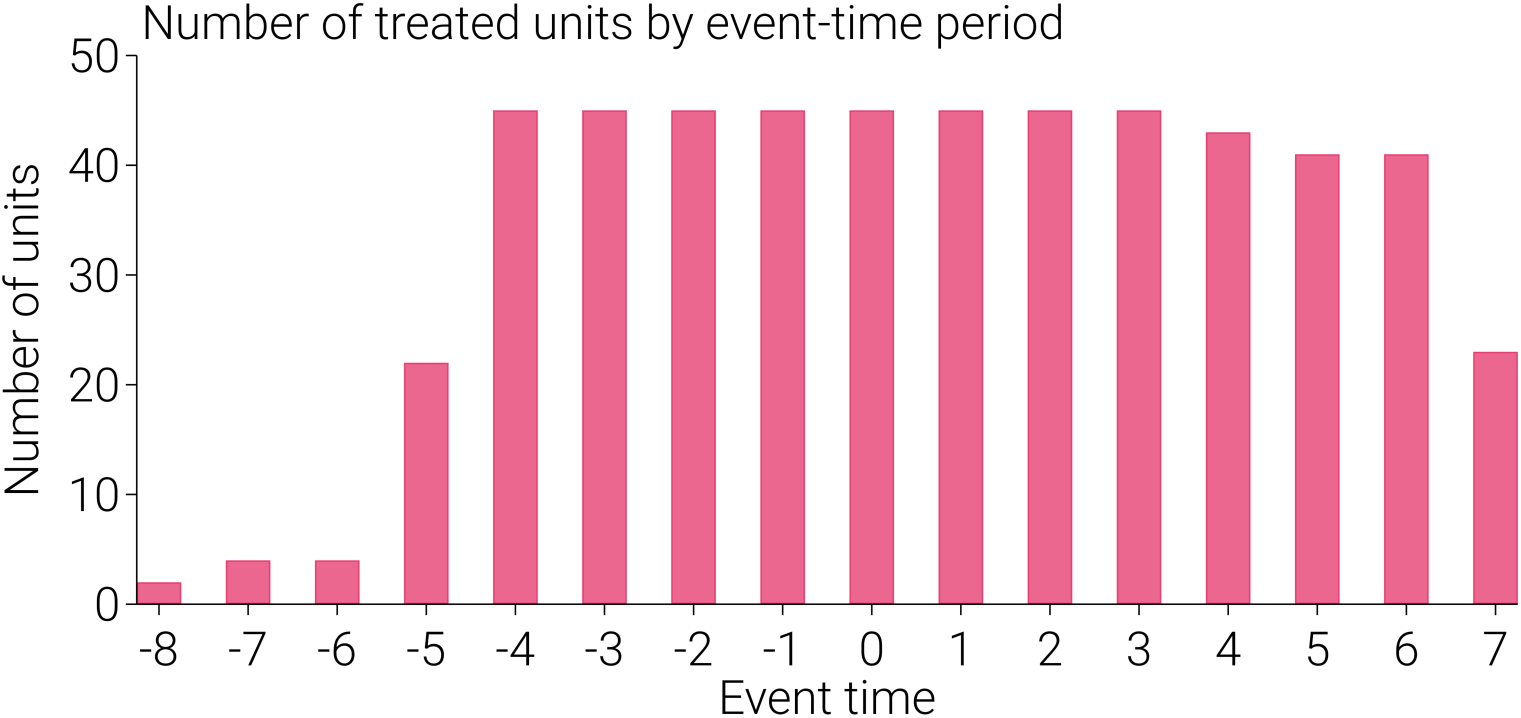
Number of Treated Counties by Event-Time Period. This figure plots the number of treated states in each event-time period from *l* = −8 to *l* = 7, corresponding to the full set of indicators that are included in the event study specifications. For clarity and due to the compositional changes in the set of treated states, the event studies do not display coefficients for periods from *l* = −8 to *l* = − 5 and from *l* = 5 to *l* = 7.

**Figure A2:**
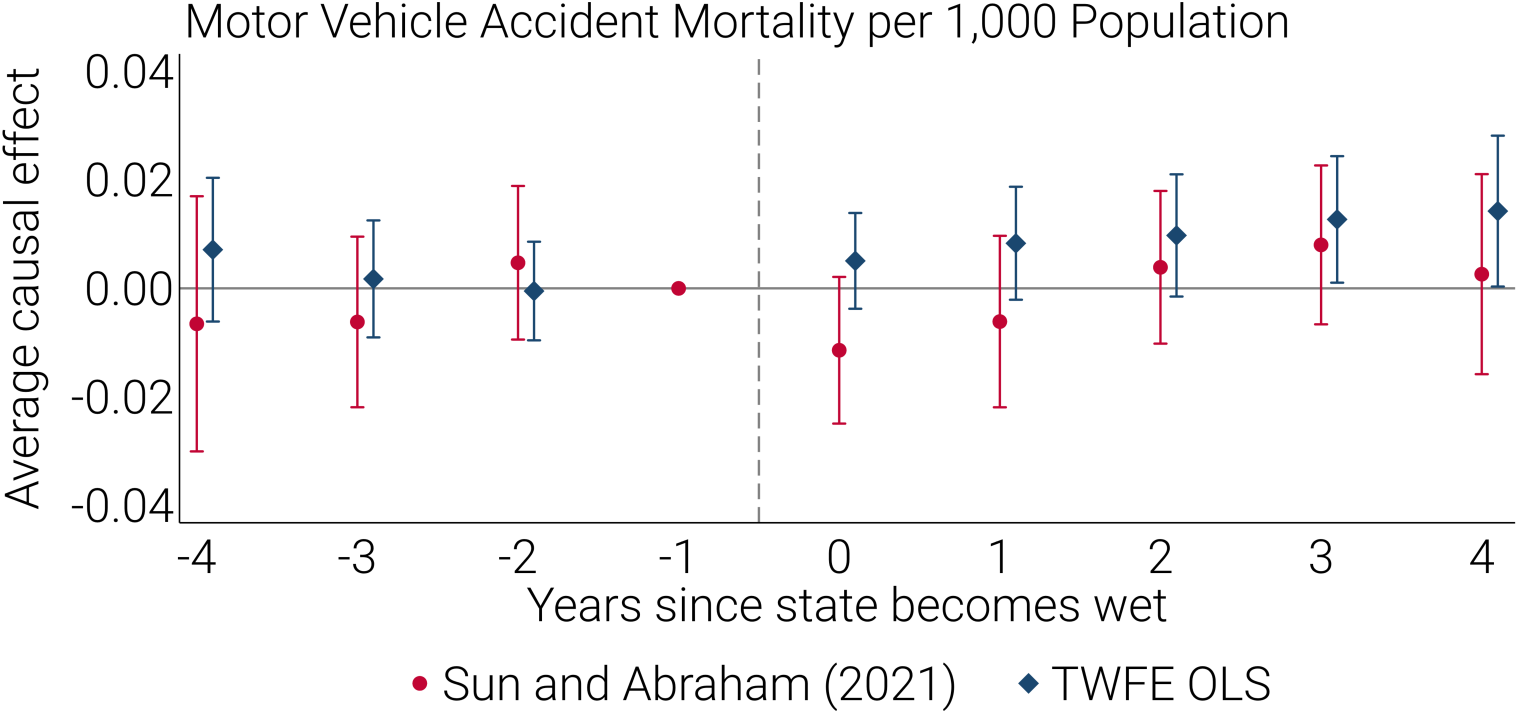
Event Study for Motor Vehicle Accidents. This figure displays an event study with motor vehicle accidents per 1,000 population as the dependent variable, using the same clustering, estimation techniques, observation weighting, sample, and specification as those in Figure 2. The sample includes the 1930 to 1941 birth cohorts. Mortality is observed between 1990 and 2004. The coefficients for event-time periods *l* = −8 to *l* = − 5 and *l* = 5 to *l* = 7 are omitted for clarity and due to the compositional changes in the set of treated states. All specifications include birth year by year of death and state of birth by year of death fixed effects. Observations are weighted by the surviving cohort size for each state of birth by year of birth by death year cell. Standard errors are clustered at the state of birth by year of birth level.

**Figure A3:**
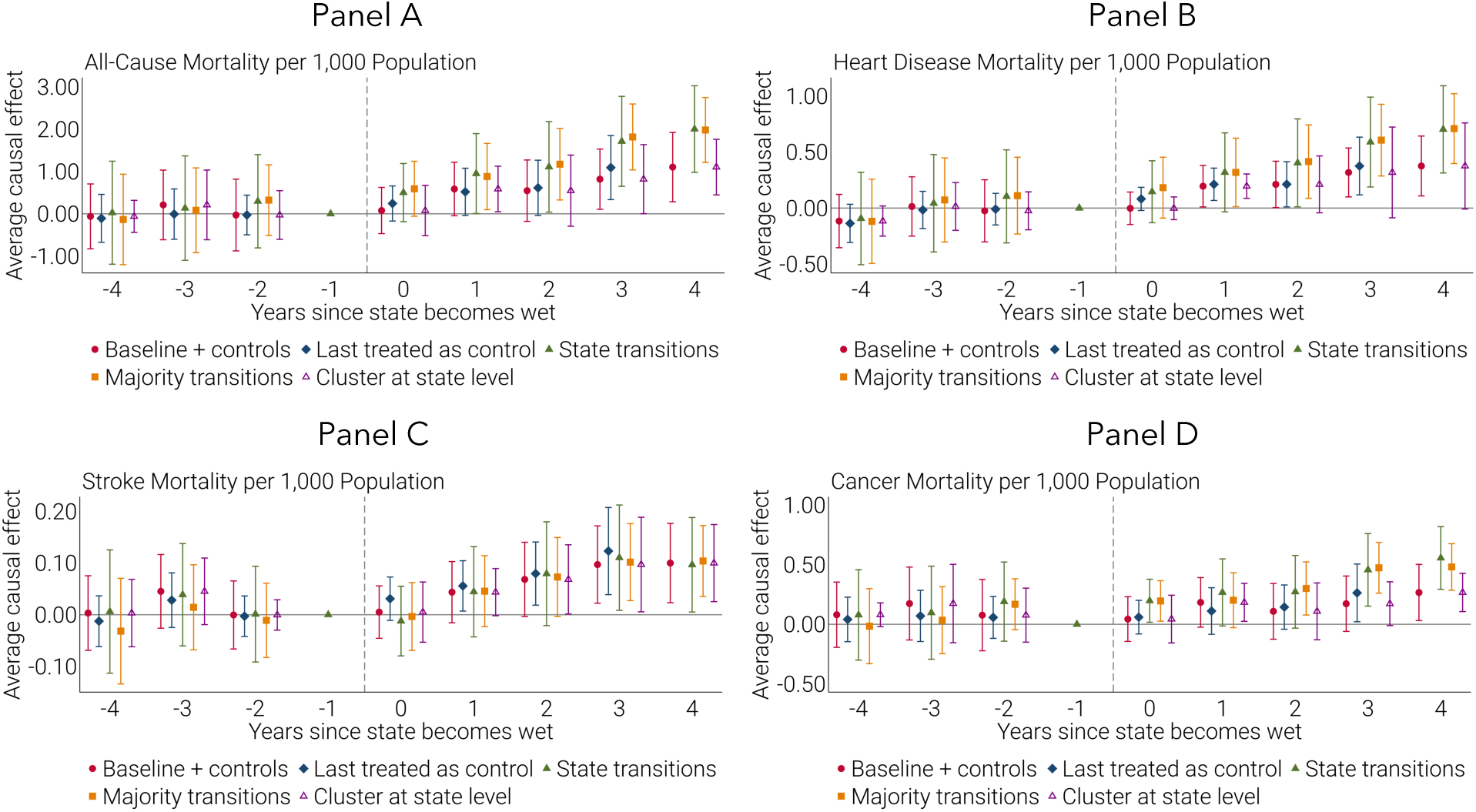
Robustness of Event Study Specifications. The panels display event studies with all-cause mortality (panel A) or cause-specific mortality per 1,000 population as the dependent variable (heart disease in panel B, stroke in panel C, and cancer in panel D). Each panel reports the event studies corresponding to the aggregated coefficients reported in columns 2 to 5 of Table 2. The sample includes the 1930 to 1941 birth cohorts. Mortality is observed between 1990 and 2004. The coefficients for event-time periods *l* = −8 to *l* = − 5 and *l* = 5 to *l* = 7 are omitted for clarity and due to the compositional changes in the set of treated states. All specifications include birth year by year of death and state of birth by year of death fixed effects. Observations are weighted by the surviving cohort size for each state of birth by year of birth by death year cell. Standard errors are clustered at the state of birth by year of birth level except for the specification labeled “Cluster at state level” which is denoted by a triangle with a purple outline.

**Table A1:** Crosswalk of ICD9 and ICD10 for Cause-Specific Mortality.

| **Cause-specific death** | **ICD9** | **ICD10** |
| --- | --- | --- |
| Heart disease | 390-8, 402, 404, 410-29 | I00-09, 11, 13, 20-51 |
| Stroke (cerebrovascular disease) | 430-434, 436-438 | I60-69 |
| Cancer | 140-208 | C00-97 |
| Motor vehicle accident | E810-25 | V02-04, 09.0, 09.2, 12-14, 19.0-19.2, 19.4-19.6, 20-79, 80.3-80.5, 81.0-81.1, 82.0-82.1, 83-86, 87.0-87.8, 88.0-88.8, 89.0, 89.2 |

ICD9 was used until 1998 while ICD10 has been used since in the Multiple Cause of Death data. Source: Anderson et al. (2001).

### Appendix B: Results from Extended Panel

**Figure B1:**
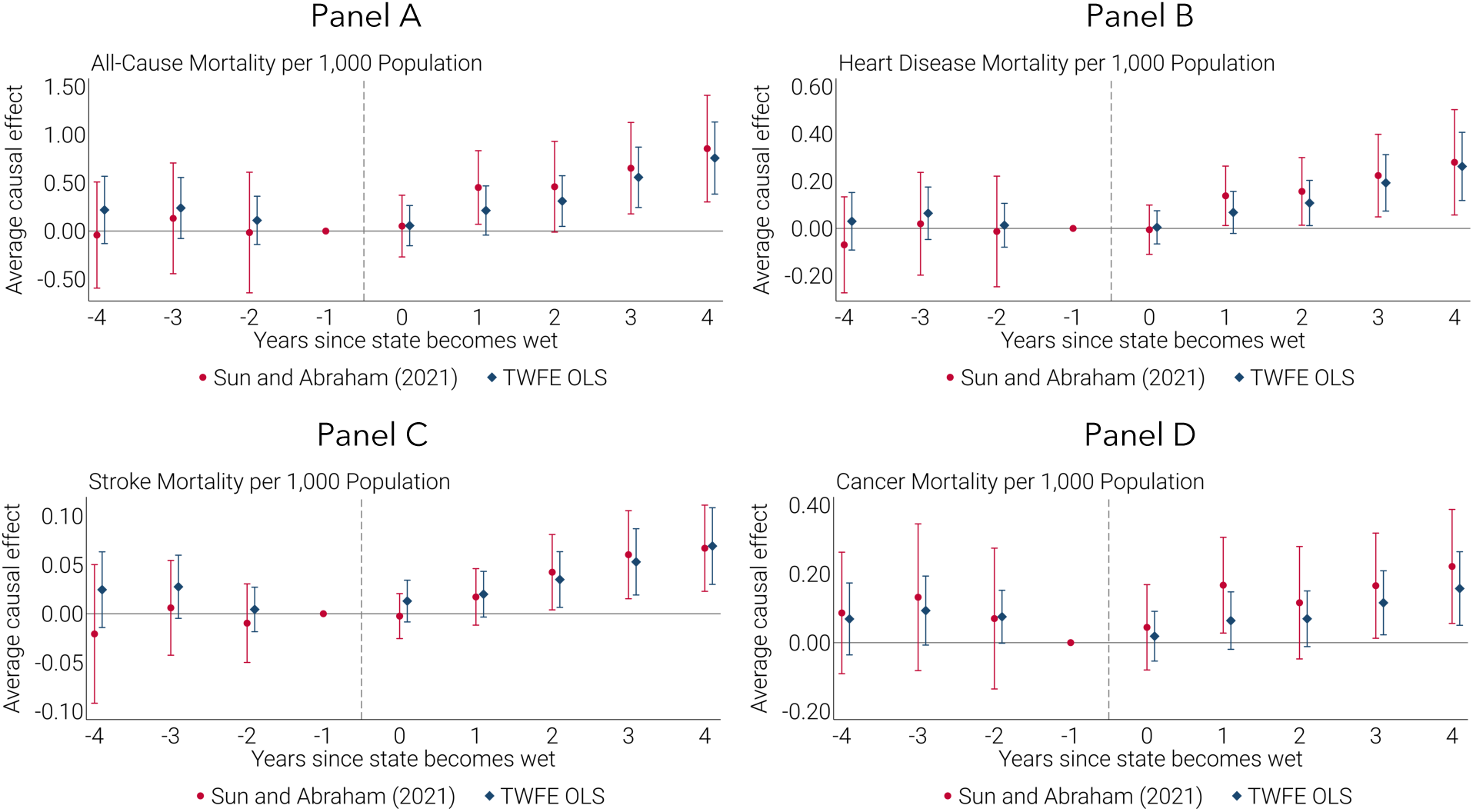
Event Studies, 1979-2004. Each panel displays event studies with all-cause mortality (panel A) or cause-specific mortality per 1,000 population as the dependent variable (heart disease in panel B, stroke in panel C, and cancer in panel D). Mortality is observed between 1979 and 2004. In all other respects, the clustering, estimation techniques, observation weighting, and specification correspond to those used in Figure 2. See the note to Appendix Figure A3 for more details.

**Table B1:** Aggregate Event Study Estimates for Cause-Specific Mortality, 1979-2004.

|  | (1) | (2) | (3) | (4) |
| --- | --- | --- | --- | --- |
|  | All-cause mortality | Heart disease | Stroke | Cancer |
| Wet status (=1) | 0.45<br>(0.20)<br>[2.26] | 0.15<br>(0.06)<br>[2.60] | 0.03<br>(0.02)<br>[1.88] | 0.14<br>(0.07)<br>[2.01] |
| Mean of Y | 13.85 | 3.86 | 0.63 | 4.89 |
| Percent effect relative to mean | 3.3 | 3.9 | 4.7 | 2.8 |
| Observations | 14,976 | 14,976 | 14,976 | 14,976 |
| Year of birth by death year FEs | X | X | X | X |
| State of birth by death year FEs | X | X | X | X |
| State by year of birth controls | X | X | X | X |

### Appendix C: Results from Continuous Treatment

**Figure C1:**
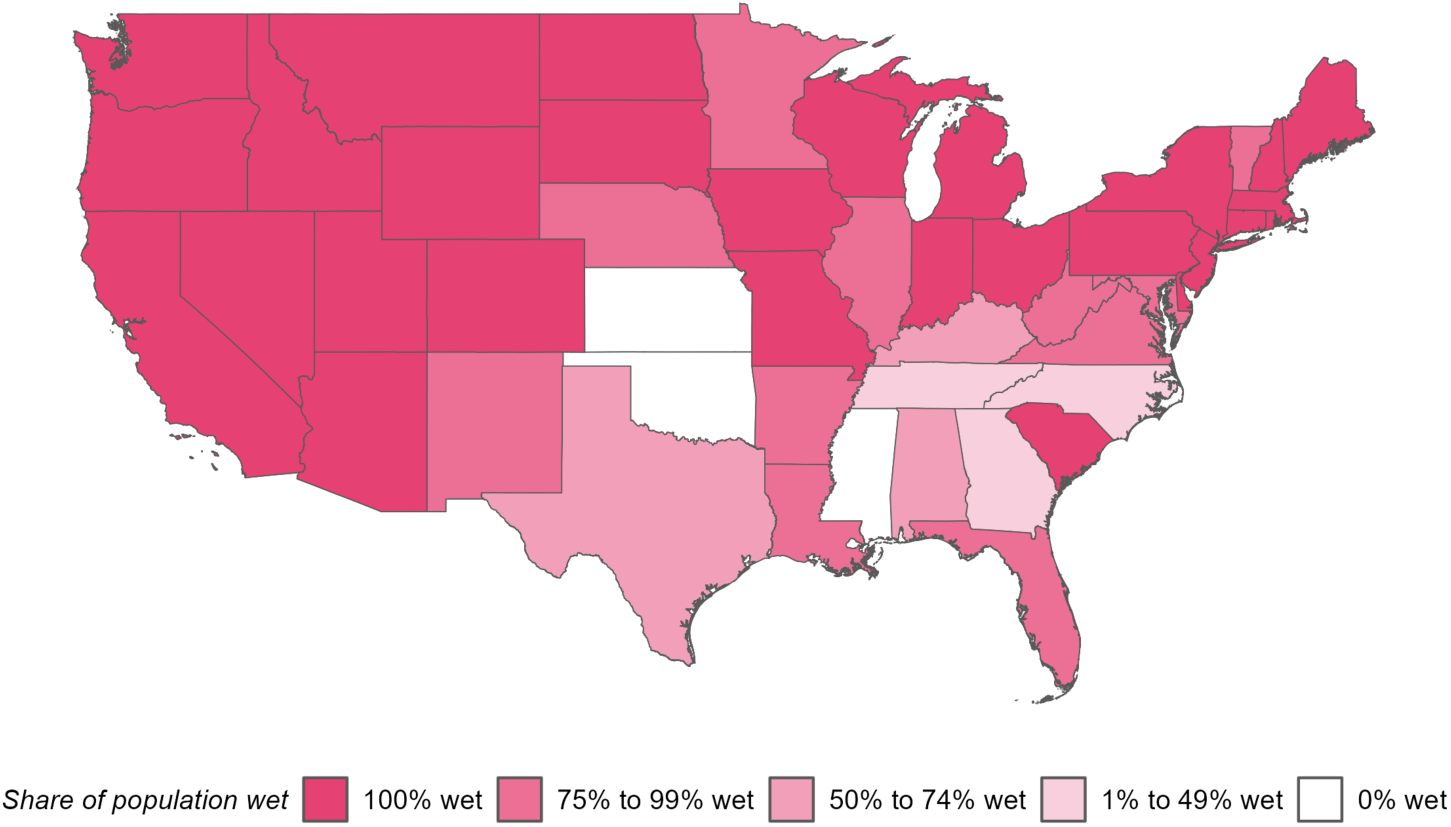
Variation in Share of States’ Population Residing in Wet Counties. This figure plots the average share of states’ population that is treated (i.e., residing in wet counties) between 1938 and 1941. In the underlying data, this share varies by year-to-year and is used as the treatment in Table C1. Map source: Manson *et al*. (2022)

**Table C1:** Difference-in-Differences Estimates for Cause-Specific Mortality with Continuous Treatment.

|  | (1) | (2) | (3) | (4) |
| --- | --- | --- | --- | --- |
|  | All-cause mortality | Heart disease | Stroke | Cancer |
| Wet status | 0.62<br>(0.16)<br>[3.76] | 0.25<br>(0.06)<br>[4.33] | 0.07<br>(0.02)<br>[4.42] | 0.08<br>(0.05)<br>[1.56] |
| Mean of Y | 18.85 | 5.12 | 0.88 | 6.72 |
| Percent effect relative to mean | 3.3 | 4.9 | 7.9 | 1.2 |
| Observations | 8,640 | 8,640 | 8,640 | 8,640 |
| Year of birth by death year FEs | X | X | X | X |
| Birth state by death year FEs | X | X | X | X |
| Birth state by year of birth controls | X | X | X | X |

### Appendix D: Results using Cumulative Mortality

**Figure D1:**
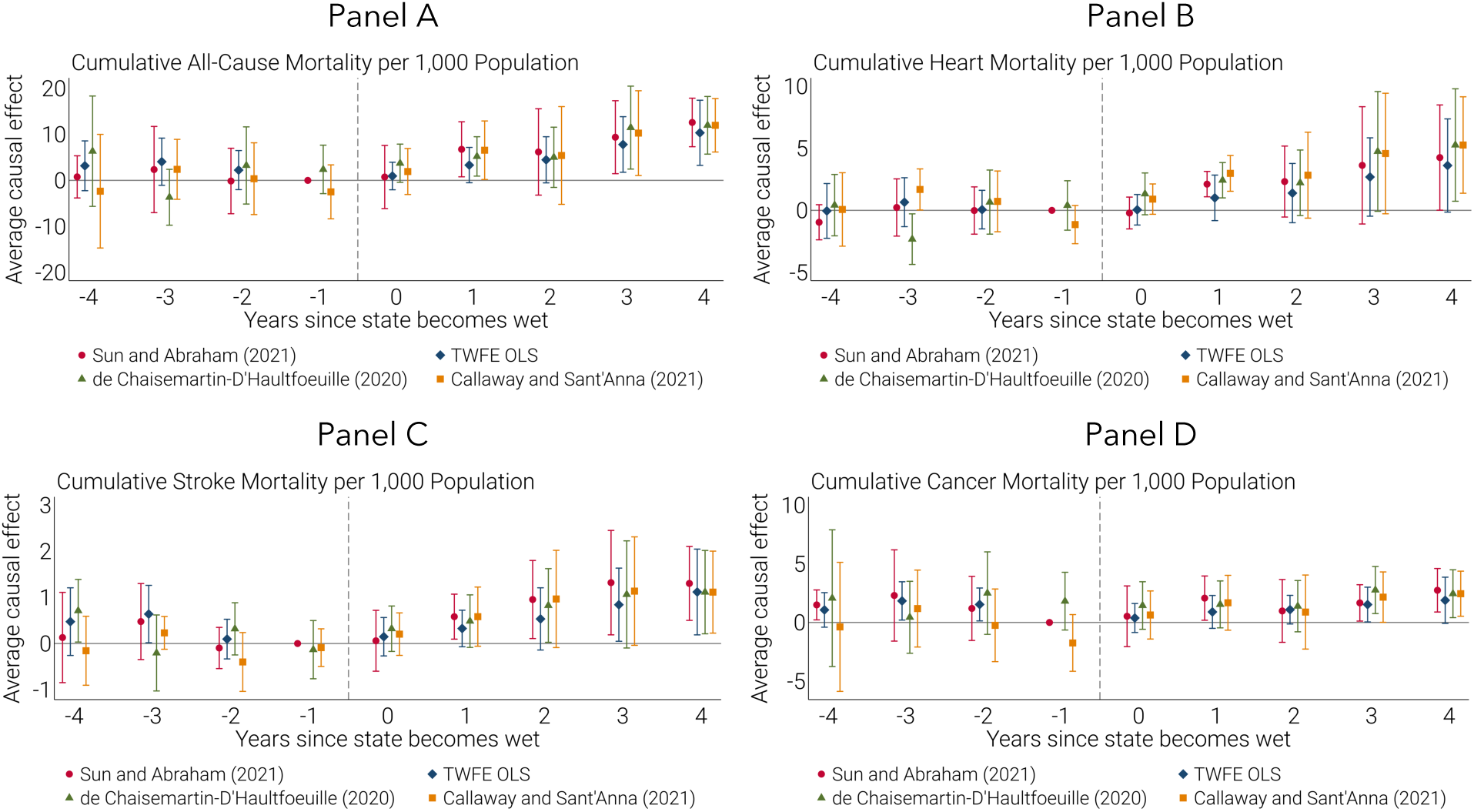
Event Studies by Cause of Death using Cumulative Mortality. Each panel displays event studies with cumulative all-cause mortality (panel A) or cumulative cause-specific mortality between 1990 and 2004 per 1,000 population in 1990 as the dependent variable (heart disease in panel B, stroke in panel C, and cancer in panel D). The unit of observation is a state of birth by year of birth cohort. The sample includes the 1930 to 1941 birth cohorts. Each panel shows estimates based on Sun and Abraham (2021) and TWFE OLS as before as well as Callaway and Sant’Anna (2021) and de Chaisemartin and D’Haultfoeuille (2020). The coefficients for event-time periods t = -8 to t = -5 and t = 5 to t = 7 are omitted for clarity. All specifications include the year of birth and state of birth fixed effects. Observations are weighted by the surviving cohort size in 1990 for each state of birth by year of birth cohort. Standard errors are clustered at the state of birth level.

**Table D1:** Aggregate Event Study Estimates using Cumulative Mortality.

|  | (1) | (2) | (3) | (4) |
| --- | --- | --- | --- | --- |
|  | All-cause mortality | Heart disease | Stroke | Cancer |
| Wet status (=1) | 6.61<br>(2.75)<br>[2.40] | 2.39<br>(1.05)<br>[2.28] | 0.73<br>(0.25)<br>[2.91] | 1.53<br>(0.87)<br>[1.77] |
| Mean of Y | 247.65 | 67.55 | 11.41 | 88.75 |
| Percent effect relative to mean | 2.7 | 3.5 | 6.4 | 1.7 |
| Observations | 576 | 576 | 576 | 576 |
| Year of birth FEs | X | X | X | X |
| Birth state FEs | X | X | X | X |
| Birth state by year of birth controls | X | X | X | X |

The unit of observation is a state of birth by year of birth cohort. The sample includes the 1930 to 1941 birth cohorts. The dependent variable is cumulative all-cause deaths per 1,000 population (Column 1) or the number of cause-specific deaths from heart disease (Column 2), stroke (Column 3), or cancer (Column 4) between 1990 and 2004 per 1,000 population in 1990. Each coefficient is the average of the Sun and Abraham (2021) event study coefficients from $l = 0$ to $l = 4$ and represents the average change in the later-life all-cause mortality rate attributable to in utero exposure to the first five years of federal prohibition's repeal. In all columns, observations are treated (i.e., the indicator for wet status equals one) if at least some alcohol sales were allowed anywhere within the state of birth in the year of birth. In all specifications, mortality rates are a function of state of birth by death year and year of birth by death year fixed effects and control for the log of per capita New Deal spending and the log of per capita real disposable personal income at the state level. Observations are weighted by the surviving cohort size in 1990 for each state of birth by year of birth cohort. All standard errors are clustered at the state of birth level. Standard errors are in parentheses; t-statistics reported below standard errors in brackets.

### Appendix E: Heterogeneity – Event Studies

**Figure E1:**
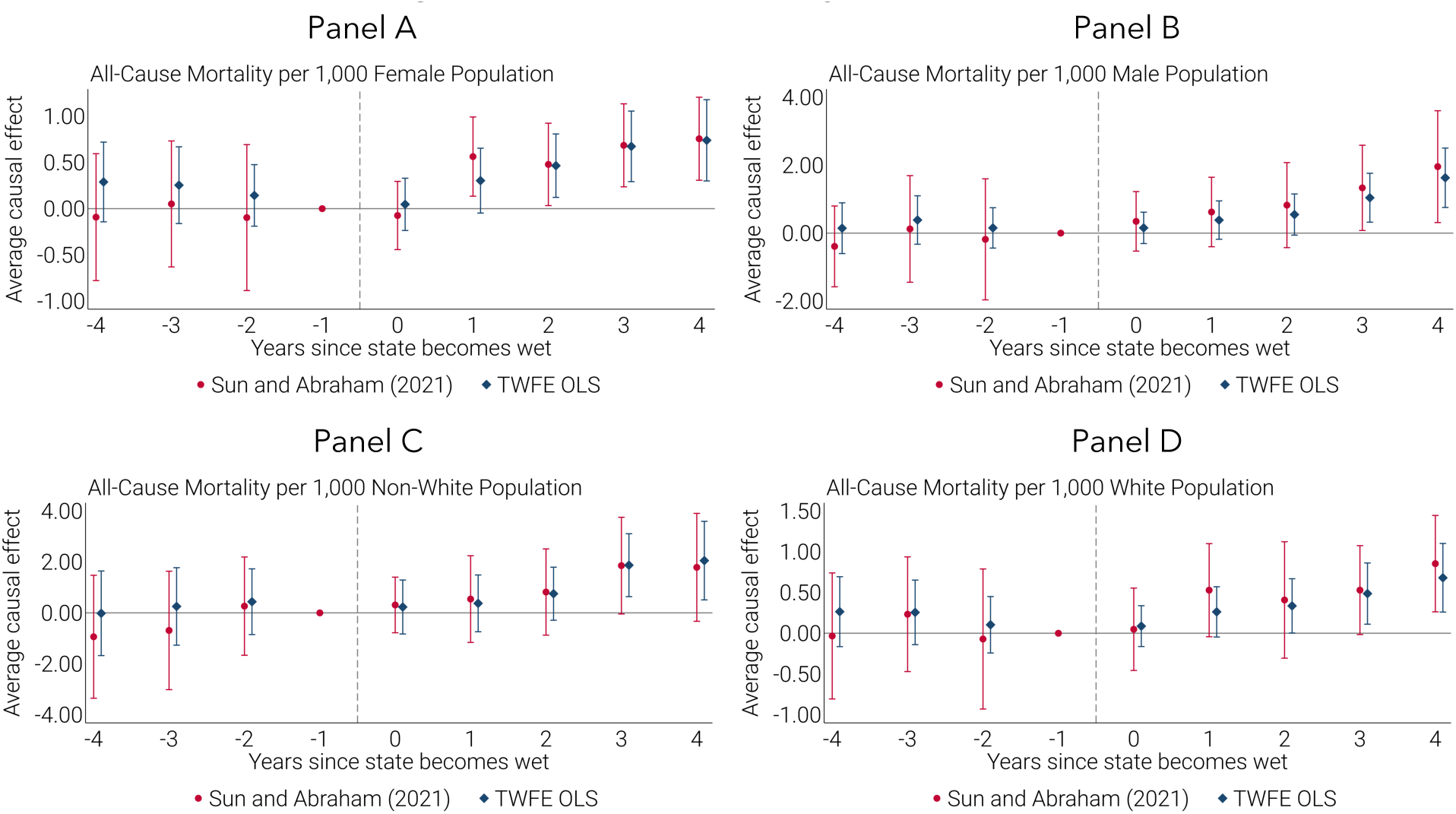
Event Studies by Sex and Race. Each panel displays event studies with all-cause mortality per 1,000 population as the dependent variable. Panels A, B, C, and D do so for females, males, non-white people, and white people, respectively. They all use the same clustering, estimation techniques, observation weighting, sample, and specification as those in Figure 2. See the note to Appendix Figure A3 for more details.

